# AI-based Prediction of Imminent Primary Stroke on Claims Data Enables Accurate Patient Stratification

**DOI:** 10.1101/2022.11.20.22282517

**Authors:** A Hilbert, D Baskan, J Rieger, C Wagner, S Sehlen, A García-Rudolph, JD Kelleher, NF Dengler, T Kossen, VI Madai, D Frey

## Abstract

**Background:** With an annual rate of 5.5 million cases, ischemic stroke is the second leading cause of death and permanent disability worldwide posing a significant medical, financial and social burden. Current approaches relax high-risk profiles of imminent stroke to mid- to long-term risk assessment, tempering the importance of immediate preventative action. Claims data may support the development of new risk prediction paradigms for better, individualized management of disease.

**Methods:** We developed a data-driven paradigm to predict personalized risk of imminent primary ischemic stroke. We used social health insurance data from northeast Germany (between 2008-2018). Stroke events were defined by the presence of an ischemic stroke ICD-10 diagnosis within the available insurance period. Controls (n=150,091) and strokes (n=53,047) were matched by age (mean=76) and insurance length (mean=3 years), resulting in a generally aged, high-risk study population.

We trained traditional and Machine Learning (ML) classifiers to predict the overall likelihood of a primary event based on 55 features including demographic parameters, ICD-10 diagnosis of diseases and dependence on care. Binary ICD-10 features were translated into temporal duration of diagnoses by counting days since the first appearance of disease in the patients’ records. We used SHAP feature importance scores for global and local explanation of model output.

**Findings:** The best ML model, Tree-boosting, yielded notably high performance with an area under the receiver operating characteristics curve of 0.91, sensitivity of 0.84 and specificity of 0.81. Long duration of hypertension, dyslipidemia and diabetes type 2 were most influential for predicting stroke while frequent dependence on care proved to mitigate stroke risk.

**Interpretation:** Our proposed data-driven ML approach provides a highly promising direction for improved and personalized prevention and management of imminent stroke, while the developed models offer direct applicability for risk stratification in the north-east German population.

**Funding:** Horizon2020 (PRECISE4Q, #777107)

## Introduction

Stroke is one of the world’s most devastating diseases putting a tremendous medical, financial and social burden on society. Both direct treatment costs and follow-up costs due to permanent disability are increasing, amounting to nearly €45 billion in 2015 in the EU^1^. Even though treatment procedures of acute stroke such as i.v. thrombolysis or mechanical thrombectomy have seen significant advances, the overall outcome after stroke is still poor. In 2017, 17.1 million disability-adjusted life years were lost in Europe due to stroke^2^.

Thus, one of the main public health goals is the prevention of stroke. By addressing major modifiable risk factors such as hypertension, diabetes mellitus and smoking, the global incidence of stroke stabilized over the last decades^3^. However, at the same time outcomes in the aged population are worsening and prevalence in the ageing population is projected to increase dramatically^3^. There are several reasons why the current prevention approaches fail to decrease the incidence of stroke. First, in current practice, prevention of stroke is primarily based on general guidelines, instead of individual identification of high-risk, which could lead to the highest impact relative to resource usage. Second, although various scores exist for the estimation of individualized combined cardio- and cerebrovascular risk, it is likely that they do not represent all stroke subpopulations well. It has been shown that stroke exhibits considerable regional variety with regards to incidence and prevalence. Thus, commonly used scores such as the Framingham score generalize poorly beyond their validation cohort and adjustments are not straightforward^4–6^. Furthermore, most scores were developed in time-constrained studies and predict risk within 5-10 years. Since many patients share general risk factors, these long-term risk profiles fail to stratify imminent high risk individuals amongst elevated risk patients. Consequently, the long-term risk prediction paradigm does not provide a proper basis for targeting immediate preventative measures required to protect individuals at a high-risk of imminent primary ischemic stroke.

Thus, to advance stroke prevention, there is an urgent need to a) employ appropriate, data-driven methods for individual prediction and flexible regional adaptation, b) define prediction paradigms and use cases for effective prevention and c) identify predictors that can discriminate between patients with similarly high risk profiles. These preventative measures targeted at very high-risk patients can further advance and accelerate the public health impact achieved in the last decades.

To this end, we identified exploitation of health insurance data through Machine Learning (ML)-based data analytics as a suitable use case. Thanks to the large variety of available features and extensive sample size for analysis, claims data represents an excellent scenario for applying ML. Machine Learning can reveal highly complex interactions between risk and protective factors and consequently could constitute a basis for better preventative treatment by facilitating personalized recommendations.

In this study, we focused on stroke prediction in a high-risk cohort, where the aforementioned potential of predictive systems could have high impact. We assessed the performance of traditional methods as well as modern ML models in prediction of imminent primary stroke in a German public health insurance cohort.

## Methods

### Data Source

Statutory Social Health Insurance (SHI) data provided by the insurer *AOK Nordost SHI fund* (AOK-NO) was used retrospectively. AOK-NO operates in the federal states of Brandenburg, Berlin and Mecklenburg-Vorpommern and covers around one quarter of insured persons. This allows us to draw conclusions for the north-east German SHI population. The original database comprises outpatient (ambulatory) and inpatient (hospital) care data on pseudonymized patient levels. In the current analysis, data within the time frame of 2008 and 2018 was included. Approximately 1,4 million people above 30 years old were insured at AOK-NO per reporting year and around 1,8 million people in total within this period.

As the base population of our study, 510.728 patients older than 30 years were pulled from the database with matched controls to each stroke patient per year. Insurance coverage in all the reporting years was not a requirement for inclusion, thus insurance times may vary.

#### Study Population

##### Stroke cohort

The base population contained 161,963 patients with continuous insurance and ambulatory and/or hospital diagnoses of ischemic stroke defined as any subclass of the ICD-10 code class I63 with all diagnosis reliabilities and types (Figure 2.a). From this, we selected patients with at least 1 hospital ischemic stroke diagnosis of main, secondary, department main, department secondary or surgical type or at least 1 ambulatory confirmed ischemic stroke - termed inclusion stroke -, resulting in 94,600 patients (Figure 2.b). Next, we excluded 29,561 patients who had an ambulatory or hospital diagnosis of cerebrovascular disease, more specifically hemorrhagic or unspecified stroke and its sequelae (I60-62.x, I64.x, I69.0-4) before the inclusion stroke, yielding 65,039 patients (Figure 2.c). Lastly, 11,992 patients were excluded who had ambulatory stroke diagnoses preceding a hospital stroke diagnosis, arriving at a final number of 53,047 stroke patients for our analysis (Figure 2.d).

**Figure 1:**
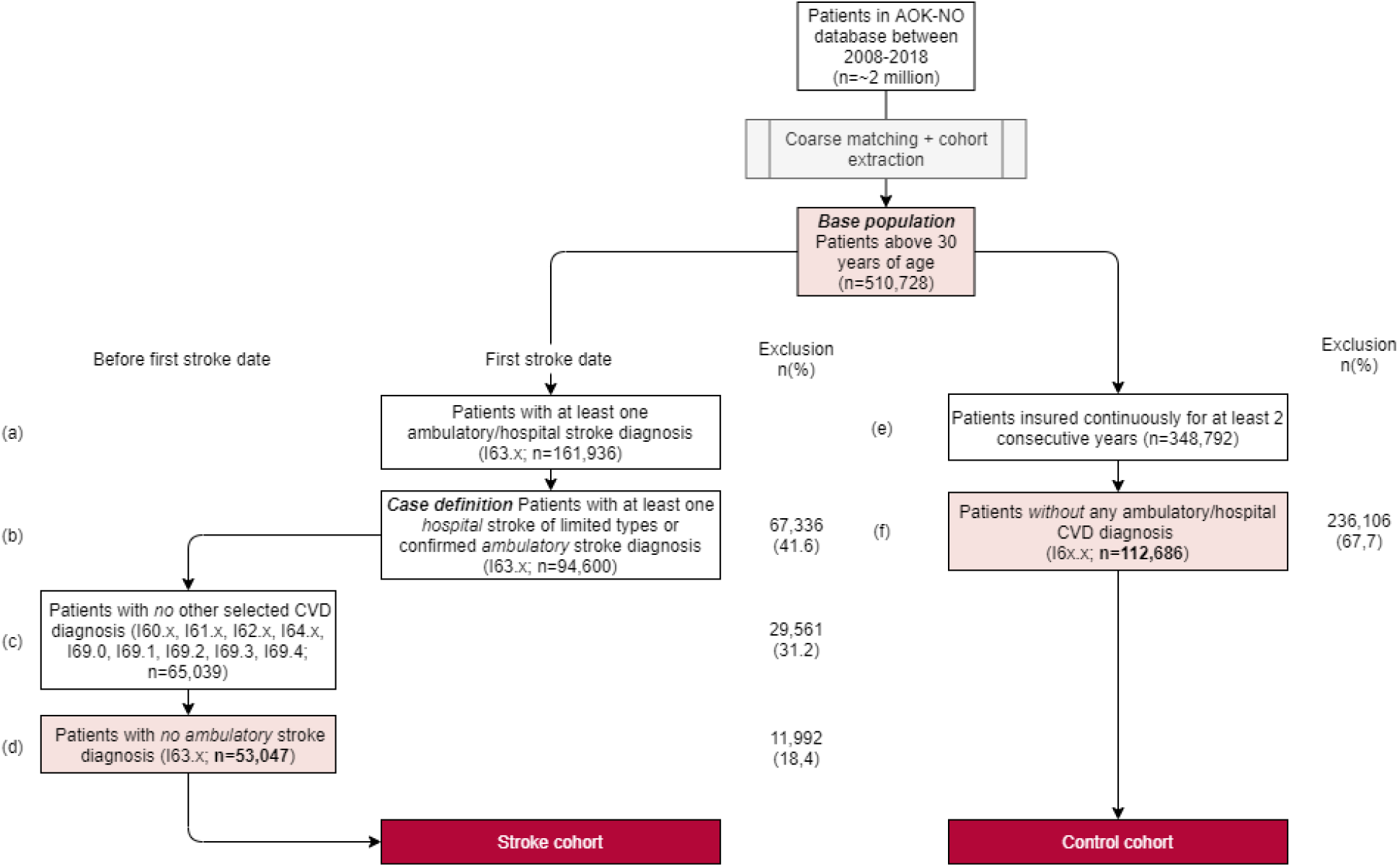
Cohort definition process

**Figure 2:**
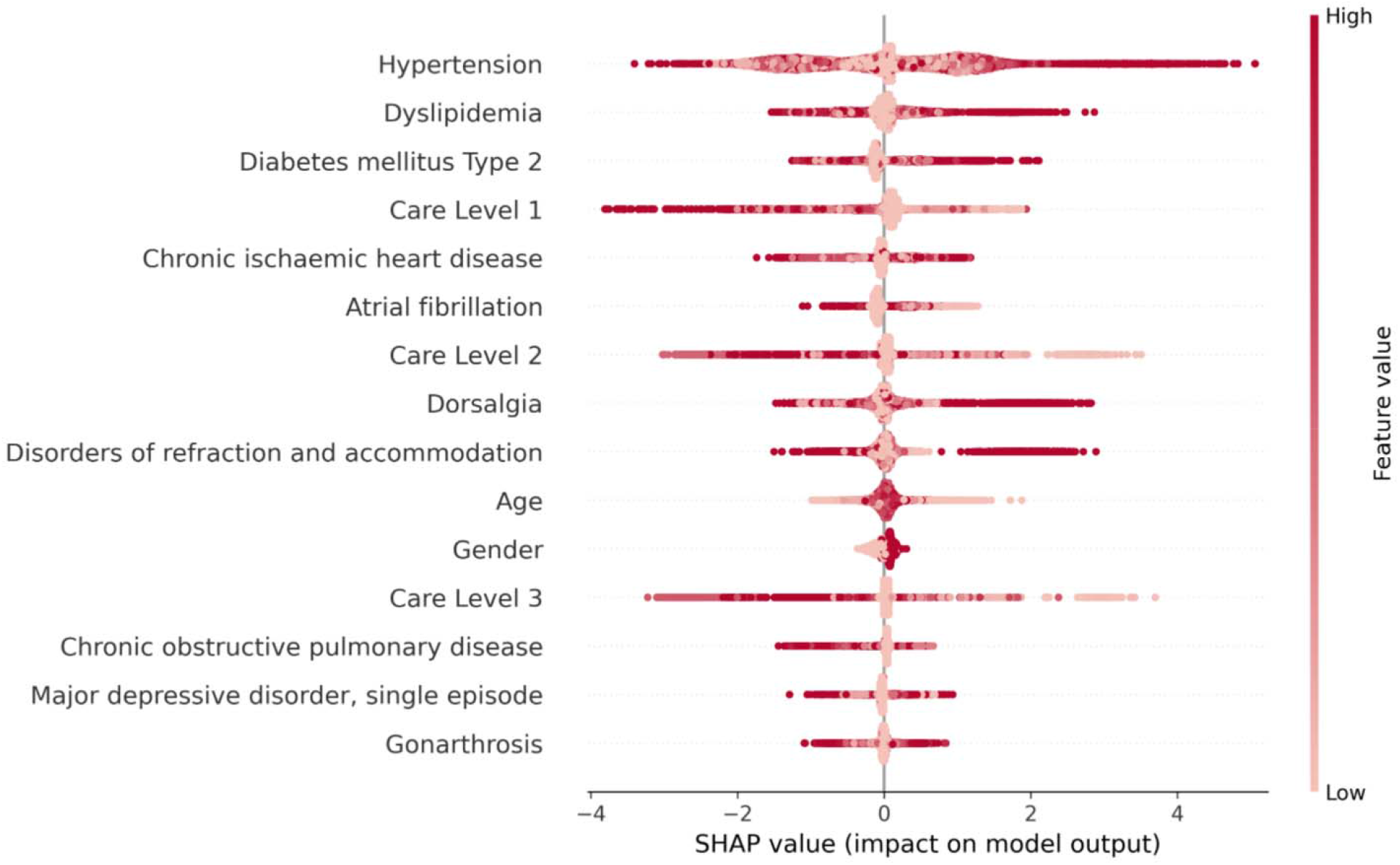
Feature importance ranking on the test set from our best model, Tree-boosting. In each row the distribution of importance of the given variable is shown. Positive SHAP values indicate impact in favour of prediction of stroke, while negative values indicate impact against prediction of stroke. All points on the graph are colored based on the true value of a current variable within possible ranges. Variables are ordered by their absolute importance aggregated from all test predictions. In general, hypertension, dyslipidemia or diabetes mellitus type 2 had the highest impact on model output and a longer record of these variables (dark red) contributed to predictions of stroke. The unusually low importance of age can be attributed to the close matching of stroke and control cohorts.

##### Control cohort

The base control population with at least 2 years of continuous insurance contained 348,792 patients (Figure 2e). We excluded 198,701 patients with any record of ambulatory and/or hospital diagnosis of a cerebrovascular disease, more specifically hemorrhagic, ischemic or unspecified stroke and its sequelae (I60-64, I69.x), resulting in a final number of 150,091 control patients for our analysis (Figure 2f).

Our final dataset included a total number of 203,138 patients; the described cohort definition process is illustrated in Figure 2.

### Risk Variables

More than 50 variables with potential to be associated with ischemic stroke were extracted. These variables included demographic variables as well as disease diagnoses given by the ICD-10 code definition and number of days of dependence on different levels of care.

Originally, ICD-10 codes up to 3 characters were considered. Variables with a prevalence of less than 5 percent were grouped with variables of the same 2 character group and discarded in case the prevalence did not reach over 5 percent after grouping. Table 1 gives a detailed overview of the distribution of the final variables included.

**Table 1:**
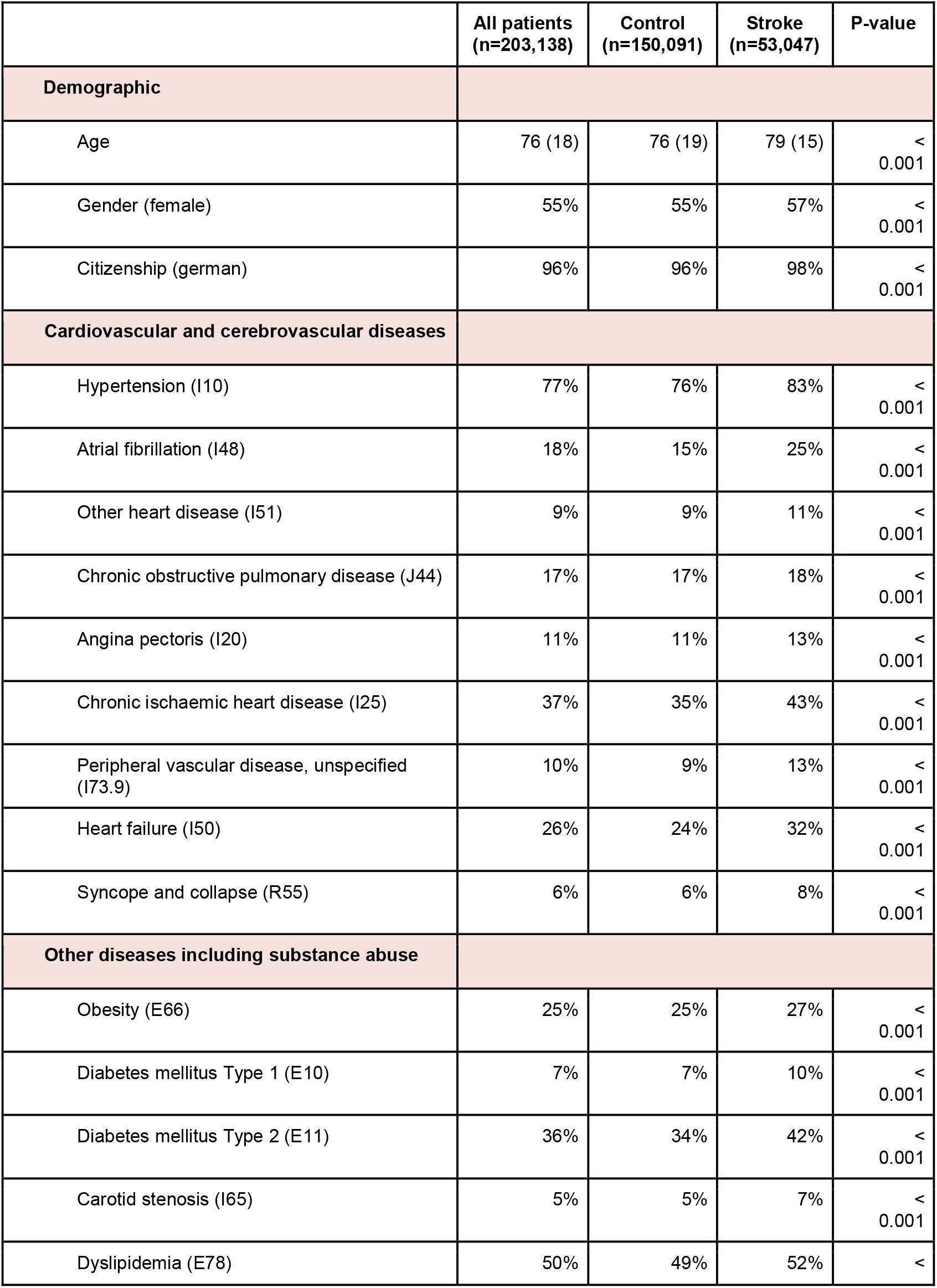
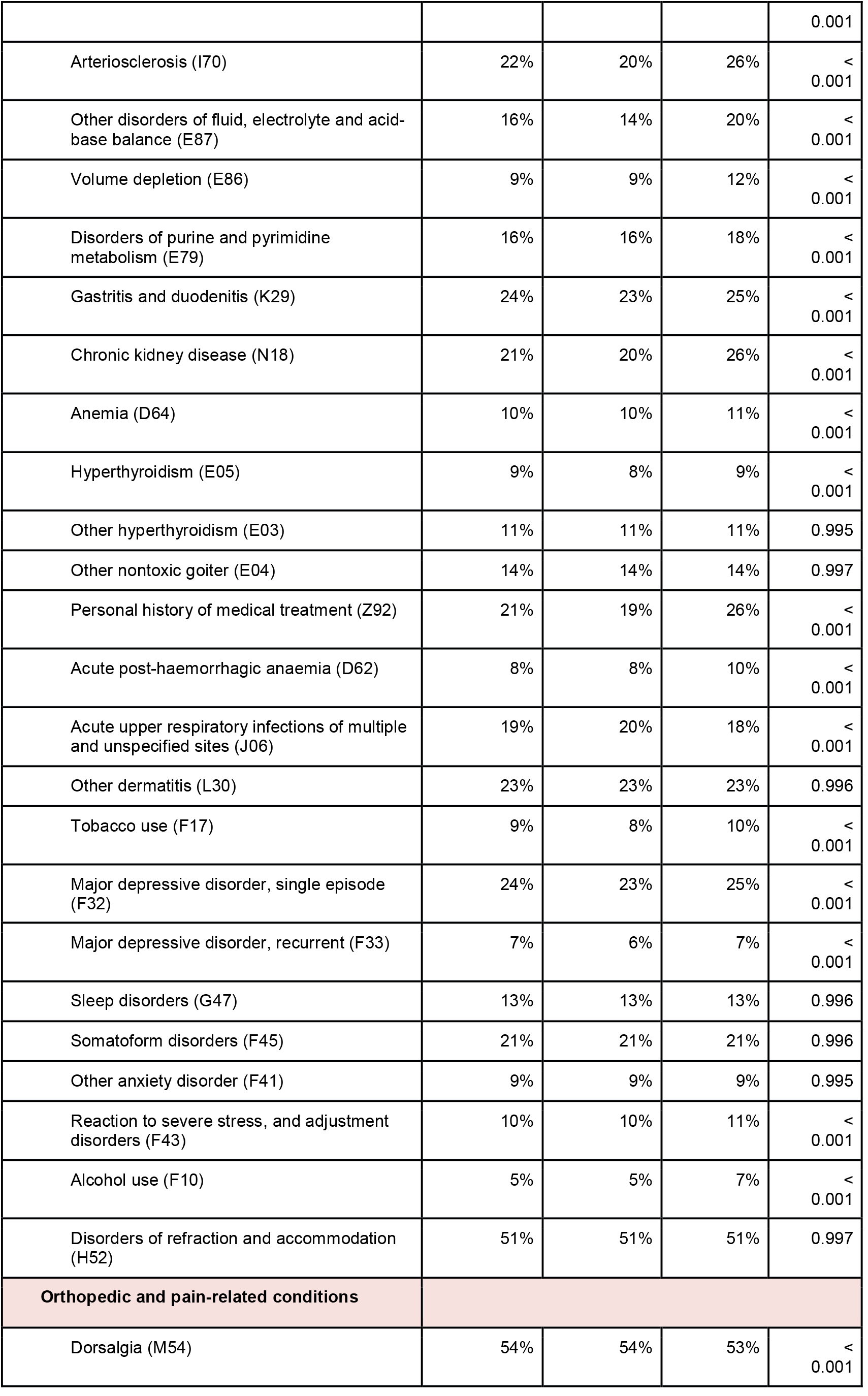
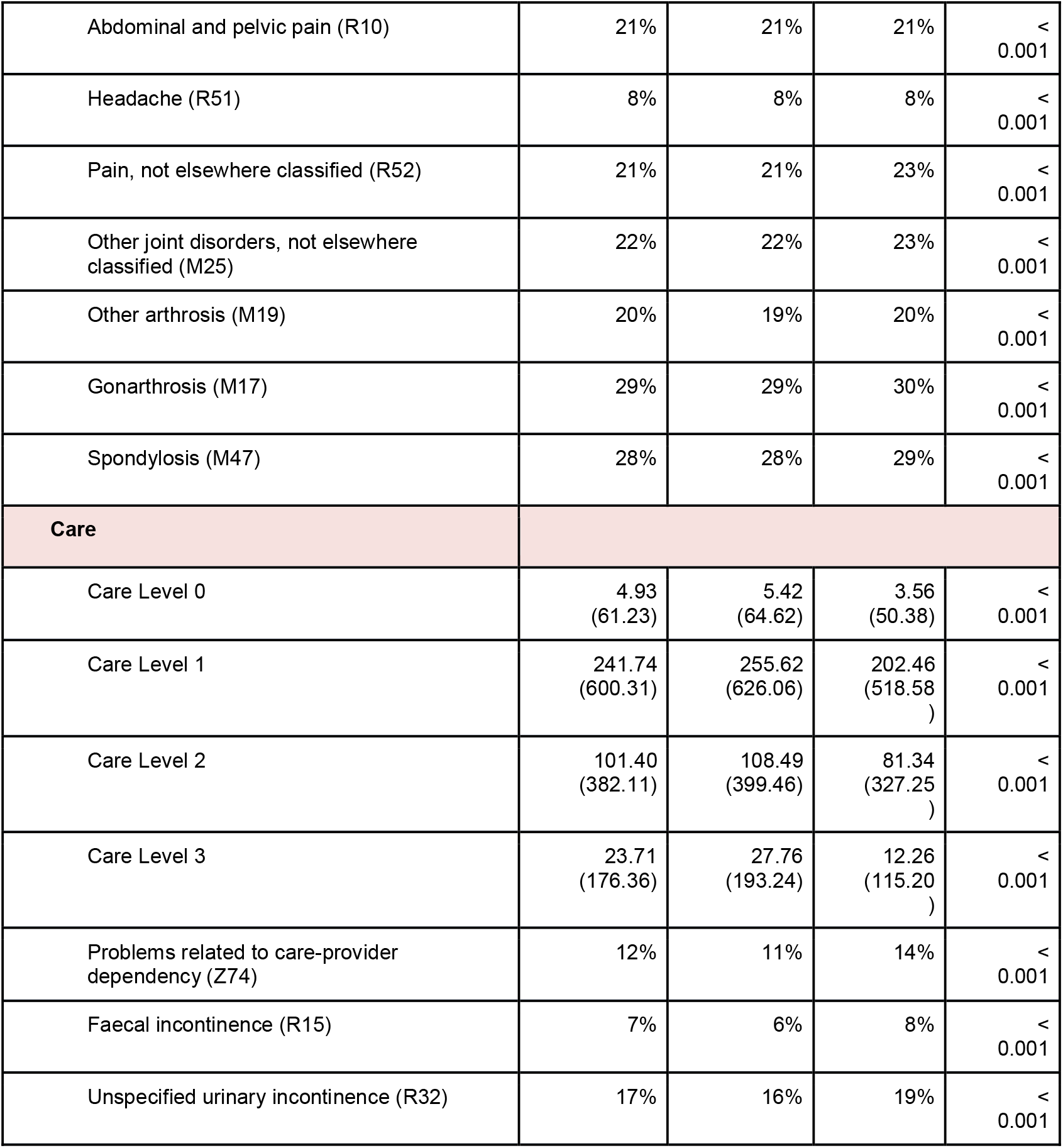
Detailed list and distribution of final variables included. Second, third, fourth columns show mean value and standard deviation in case of continuous variables (age and care levels) and ratio of the categories in case of categorical variables in the complete study population as well as the control and stroke cohorts respectively. Fifth column gives the statistical significance of the difference between the control and stroke cohorts for each variable in terms of p-values derived from Pearson’s Chi-squared statistics for categorical and two-tailed t-test for continuous variables.

Even though in Germany care levels were substituted by care grades as of 2017, we chose to utilize care levels as most of the patients’ records originated prior to 2017. Patient records containing care grades were thus transferred to their approximate care levels (Care grades<3→ Care level 1, Care grade 3 → Care level 2, Care grades>3 → Care level 3). Care levels are based on the amount of time a patient requires assistance such as personal hygiene, nutrition and mobility on a daily basis, thus higher means more severe.

### Feature Representation

In contrast to longitudinal study settings, we defined the event date (stroke vs. no-stroke) and collection period individually for each patient and collected the variables from the most recent records available in those periods. This creates our desired imminent prediction paradigm, where a prediction of stroke reflects the imminent and overall likelihood for a future primary event of an individual. For our analysis we considered only patients who were insured continuously. In the insurance period we tolerated short insurance breaks with a maximal of 15 days and included those days in the insurance period (short breaks originate primarily from administrative errors).

Next, we created a temporal representation of ICD-10 features, describing the duration of diagnosed diseases. Binary ICD-10 classes were translated into the persistence of a given condition defined by the days elapsed since its first occurrence in the patient’s record till the event date. Demographic features were left intact and care features were given in days originally in the dataset.

The extracted features of 203,138 patients were used to construct the final dataset for model development. We employed close adjustment and matching of individual time periods between controls and stroke to eliminate any temporal mismatch and bias. Hence, the final mean age and insurance length were 76 and 3 respectively for both control and stroke cohorts. Detailed description of definition of collection periods as well as temporal matching procedure can be found in the Appendix.

### Outcome

The objective of the prevention model is to predict the imminent occurrence of primary acute ischemic stroke for individual patients defined as a cerebrovascular event falling into any subclass of the ICD-10 code class I63. Due to the imminent prediction paradigm, primary events at any time during the available insurance period for patients were taken into account. Consecutive events were not considered in the analysis.

### Machine Learning Algorithms

#### Machine Learning Framework

To train classifiers for stroke prediction, a publicly accessible ML framework developed by the authors was adopted and utilized. The framework has been used and the technical implementation has been described in open access publications previously^7^. The program code is written in Python using standard ML libraries and is available on Github (https://github.com/prediction2020/explainable-predictive-models).

#### Applied Algorithms

We utilized all four ML algorithms from the framework, namely Tree-boosting, Support Vector Machine (SVM), Naive Bayes (NB) and Multilayer Perceptron (MLP) to provide a comprehensive coverage of ML methods of various nature. Traditional techniques were represented by Logistic regression (LR) and its regularized variants: Lasso and ElasticNet. We estimated multicollinearity of the features using the variance inflation factor (VIF) to prevent the use of redundant features and thus potential negative impact on results as well as ensure a valid interpretation of individual feature importance^8^.

#### Model Training and Validation

The data comprising the extracted variables and outcomes were randomly split into training and test sets in a corresponding 4:1 ratio. Continuous features - all, except citizenship and gender - were standardized to zero-mean and unit variance based on training set statistics. Models were trained and best parameters were selected using 4-fold cross-validation over an extensive grid of hyperparameters; details in the Appendix. Training of all models gained significant stability due to the extensive dataset, thus we did not experience any significant variation in results when repeating this procedure with different training, validation and test sets.

#### Performance Assessment

Model performance was primarily assessed by Area Under the Receiver Operating Characteristic (AUROC) as one of the most robust and used metrics, as well as Sensitivity and Specificity to provide a more interpretable assessment of the rate of true positive and negative predictions. Additionally, accuracy, balanced class accuracy, precision, f1 score and negative predictive value measures for each model are included in the Appendix. Best hyperparameters were selected with respect to AUROC performance on the validation set.

#### Interpretability Assessment

Explainability and transparency are key concepts in translation of AI into the medical domain. A common practice augments the model’s predictions with so-called feature importance scores, that determine the contribution of the input features in the model’s prediction. We used SHapley Additive exPlanation (SHAP) scores to rate the importance of included variables for all models. SHAP is one of the most used methods in the field because of its distinct advantages such as providing a unified solution for local and global feature importance. The SHAP score of a certain variable corresponds to the marginal contribution of that variable in the model’s prediction. Used for interpretation in a single case - local importance -, it reveals the individual risk and protective factors in the specific patient’s record that the model based its prediction on. While used in an analysis over multiple samples, it provides insights into global characteristics through the lenses of the trained model. More detailed explanation of the technique can be found in ^9^. We show global importance ranking using the SHAP summary plot, and give an example for interpretation of an individual prediction through the SHAP decision plot.

### Reporting guidelines

We followed the TRIPOD reporting guidelines the corresponding checklist can be found in the Appendix.

## Results

In the multicollinearity analysis, features in every scenario demonstrated negligible relation to each other, with VIFs below 4. With regard to guidelines in ^8^ we did not recognize a harmful level of multicollinearity in any of the variable sets hence no features were eliminated.

### Model Performance

The specific AUROC, Sensitivity and Specificity measures for traditional and ML methods are shown in Table 2. Results for additional performance measures as well as additional feature representations are provided in the Appendix.

**Table 2:** Performance of all traditional and ML models on test set. Tree-boosting significantly outperformed all other methods in terms of AUROC, sensitivity and specificity.

| Models | AUROC | Sensitivity | Specificity |
| --- | --- | --- | --- |
| LR | 0.59 | 0.58 | 0.55 |
| Lasso | 0.59 | 0.58 | 0.55 |
| ElasticNet | 0.59 | 0.53 | 0.59 |
| SVM | 0.56 | 0.57 | 0.56 |
| Naive Bayes | 0.56 | 0.36 | 0.69 |
| MLP | 0.68 | 0.69 | 0.52 |
| Tree-boosting | **0.91** | **0.84** | **0.81** |

Amongst the ML methods, Tree-boosting achieved the highest performance, specifically an AUROC of 0.91, sensitivity of 0.84 and specificity of 0.81. All traditional methods as well as SVM and Naïve Bayes showed significantly lower performance.

Tree-boosting, proved superior in all considered feature representations (see Appendix) and was able to achieve notably high prediction performance with respect to all measures by exploiting temporal representation of ICD10 features. Other models remained at a moderate accuracy.

### Feature Importance Ranking

Feature importance values were computed using predictions of the best ML model. Figure 3 shows a global aggregation of feature importances for the whole test set, while Figure 4 depicts interpretation of an individual prediction. Positive SHAP values indicate the variable’s impact in favour of prediction of stroke (risk factor), negative values indicate impact against prediction of stroke (protective factor); displayed on the x-axis of Figure 3 and on the corresponding side of the vertical line in Figure 4. In Figure 3, the colors represent the true value of a given variable. For example, this translates to low values (bright) representing shorter recorded persistence of ICD10 features or fewer days in care and male gender, while high values (dark) represent the corresponding counterparts. Variables are ordered by absolute impact on model predictions in both figures.

**Figure 3:**
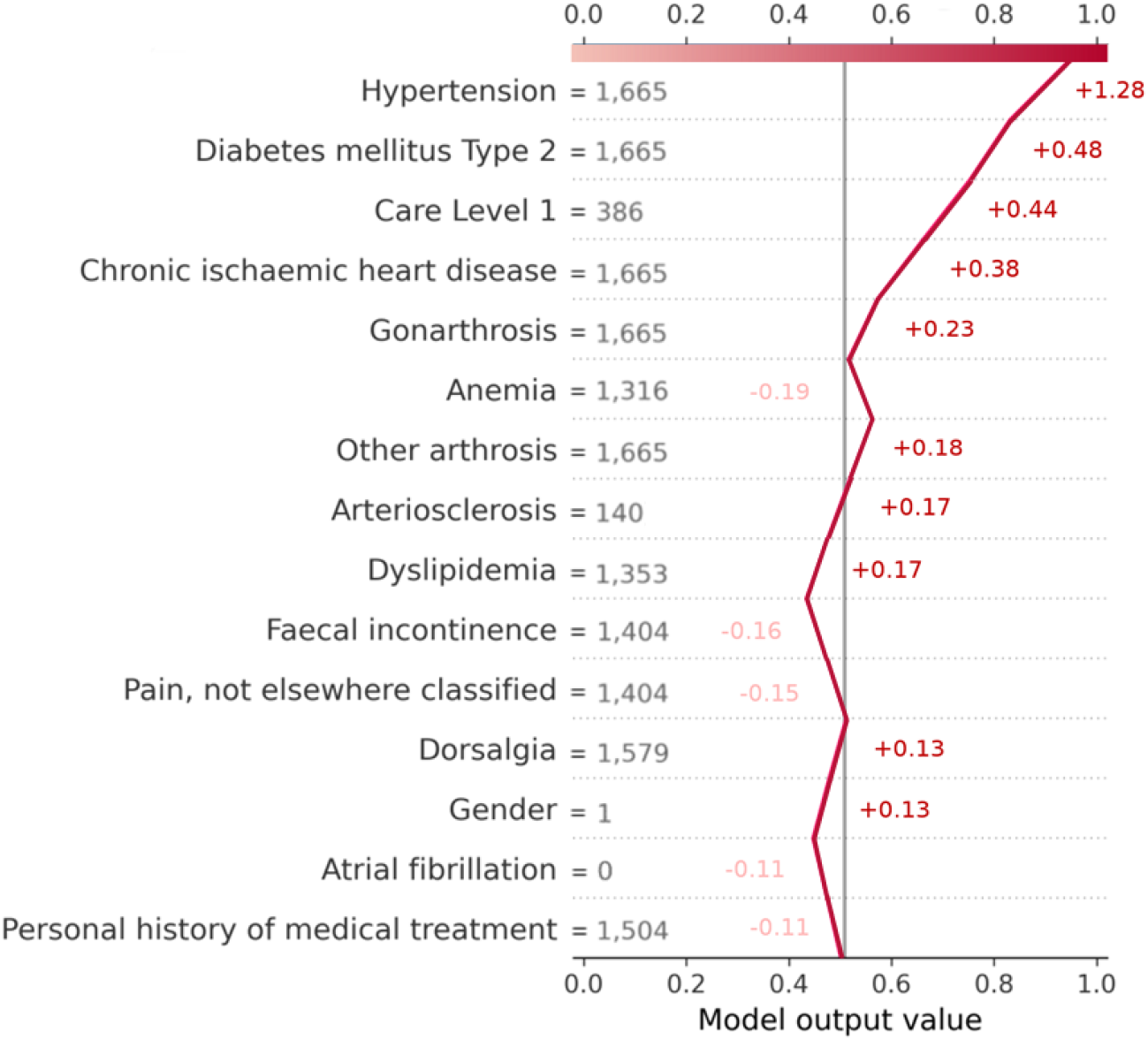
Decision plot, 84 year old female patient, predicted likelihood of stroke 0.95. Features are ordered by their average importance values. Lines going to the right imply contribution towards prediction of stroke, lines going to the left imply the opposite. Model output above 0.5 is compiled as prediction of stroke.

Globally, the most important predictive features for primary stroke occurrence were: long record of hypertension, dyslipidemia or diabetes mellitus type 2 and few days of dependence on care level 1. Moreover, the shown example case reveals the model’s reasoning for a highly certain prediction. The true values of a feature in the current patient’s record is shown next to each variable in days. Beyond the significant impact of general risk factors, the graph highlights a few protective features such as long history of anemia or no history of atrial fibrillation.

## Discussion

In the current study, we analyzed claims data of more than 200,000 high-risk patients from North-East Germany and were able to achieve a notably high prediction performance of a primary ischemic stroke event, namely an AUROC 0.91 (Sensitivity 0.84, Specificity 0.81). We showed the superiority of ML methods in comparison to traditional predictive models in modelling complex feature interactions as well as incorporation of temporal information.

Despite the reportedly reduced stroke incidence, projections forecast a continuously growing absolute stroke burden over the next 30 years in Europe^10^. This calls for imperative efforts in disease management and reassessment of research directions and paradigms supporting practical implementation of prevention strategies. Standardized risk scores such as Framingham^11^ and CHA2DS2–VASc^12^ (for Atrial Fibrillation patients) have been historically developed in longitudinal, population-based studies. Despite the poor translation of this paradigm to personalized risk, similar studies are being performed^13–17^ and a significant amount of work focuses on finding the better models for risk estimation in such datasets^18–22^. Besides, claims and electronic health record data have been exploited to retrospectively recreate and simulate a time-constrained study setting by limiting extraction of data to arbitrary time frames^23–25^. In some cases time frames correspond to annual/biannual national health check-ups, in others it follows the arbitrary 5-10 year risk estimation paradigm.

Even though prediction performances have increased over the years, we argue the practical translation of models in this long-term risk prediction paradigm to prevention is limited by design.

The key to prevent a potential stroke is action; a decision based on proper understanding of the risk and to be rigorously carried out by those concerned. The presentation of the risk as worsening conditions in 5 or 10 years relaxes the severity of a likely incidence, potentially delaying preventative action. Even though there have been several attempts to predict overall risk of stroke, the employed patient cohorts and performances are rather limited^26–28^.

Therefore, we refrained from the common long-term risk paradigm to emphasize the importance of instant intervention and showed its potential for effective preventative modelling on an extensive cohort. Translation of our results show that our best model could have successfully identified and warned 84 out of 100 high risk patients who developed a primary stroke and 81 out of 100 “benign” high risk patients from our test cohort.

Despite having great potential, often ML models fail to meet the expectations due to unclear use cases and biased data. Here, we demonstrated an example where ML could play its strength by clearly defining a high-impact use case for a specifically outlined population. This serves as a blueprint for other research groups to adapt our proposed paradigm and to replicate our results for geographically constrained population data.

In our corresponding analysis of feature importance we found all well known risk factors of stroke ranked high, confirming the importance of close monitoring also in an aged population. In contrast to previous studies, age did not play a prominent role and showed limited importance in predictions. This can be explained by the combined effect of 1) an age-matched cohort and 2) the fact that in such a diseased cohort age behaves as a compound reflection of accumulated comorbidities or dependence on care. Including all compounds in the analysis, age as an isolated feature becomes negligible, especially in case of a closely matched population. Interestingly, the number of days spent in care seems to play a surprising role; fewer days supported prediction of stroke in each level. Due to our generally higher risk cohort, this result could reflect that stroke risk might be mitigated for those relying on care more frequently merely by regular monitoring.

Furthermore, we showed a direct solution to analyze individual predictions through the SHAP decision plot. This solution facilitates the thorough understanding of present risk and potential protective factors and arms doctors to derive the best preventative strategy for the individual. Our highly accurate models and the suggested feature importance frameworks could be directly compiled into a software solution to advance prevention in the studied region.

To improve management of disease, proper deployment and distribution in the national healthcare system is as crucial as high performance. Here, we see two potential use cases. First, care providers, who are in direct contact with patients could routinely run predictions for preventative stroke screening with our solution deployed in clinical systems. In this case, calling for consecutive examination of certain risk sources or prescribing regular check-ups is immediately possible. Second, insurance companies, who host and maintain health records of patients could integrate predictive algorithms into their infrastructure and refer patients to disease management programs through stratification. Here, effectiveness greatly depends on an established chain of intersectoral connections between insurance companies, healthcare providers and patients. Even though, most of the features used by the presented models do not change rapidly and are rather chronic conditions, predictions might be less applicable in a real-time setting in case of greater data administration overhead.

The studied cohort comprises generally diseased patients – seen e.g. by the 5% higher prevalence of hypertension in controls compared to the 2008-11 German reference^29^ –, further challenging stratification of imminent high risk profiles. The translation of binary presence of disease features into numerical values of persistence gives rise to a higher degree of potential variations in the data and thus enables the identification of more complex decision boundaries. In our analysis, we demonstrated an ML based model that could make use of this potential and improve imminent stroke prediction.

Moreover, in contrast to existing manual scores estimating stroke risk, our employed features and data preparation are universal and not limited to stroke. This means, our paradigm have good potential to apply to prediction of other diseases, where chronic conditions act as catalysts in triggering adverse events.

Our study has several recognized limitations. Firstly, due to the exclusive representation of data from North-east Germany, external validation is warranted and developed models should be fine-tuned to be applicable in other regions where prediction performance and overall importance of features might change. Secondly, we developed our models in a retrospective dataset retrieved from an insurance database. The practical, deployed usage and real-world performance of a prediction model, however, is highly dependent on the data distribution presented during the model’s lifecycle. To prevent degradation of performance and model staleness, continuous monitoring and statistical analysis of incoming data compared to the training data is warranted in a real-world setting as outlined in previous research^29^. Lastly, care grades were translated into care levels, an older notation in the German health system. This means that execution of this translation step is required a-priori to retrieve future predictions, however models can also be adapted to the new notation through minor fine-tuning.

In conclusion, we tackled imminent stroke prediction from a novel angle and developed a highly accurate ML model that could enable effective screening and advanced individualized management of ischemic stroke.

## Data Availability

Models produced in the present study are available upon reasonable request to the authors.

## Contributions

AH, DF, VM conceptualized and designed the study. DB, JR reviewed the literature. AH, DB, CW, JR verified the data. AH, DB, JR and CW conducted the preliminary data analysis and extraction. AH and DB developed the final models while CW contributed to model development. AH, DF interpreted results. AH wrote the first draft, all other co-authors contributed to subsequent versions and AH finalized the manuscript. All authors contributed to the critical revision and read and approved the final manuscript.

The corresponding author (DF) attests that all listed authors meet authorship criteria and that no others meeting the criteria have been omitted. AH and DF had final responsibility for the decision to submit for publication. All authors had full access to all the data in the study and accepted responsibility for the decision to submit for publication.

