## Supplemental Material for "AI-based Prediction of Imminent Primary Stroke on Claims Data Enables Accurate Patient Stratification"

### Appendix

Feature collection periods

In contrast to longitudinal study settings, no general event date was determined for the patients. The event date was individual for each patient and its identification strategy differed for stroke patients and controls. In case of stroke patients, the event date was set to the patient’s first hospital admission date with ischemic stroke diagnosis. In case of controls, an slack period was defined, starting 1 year before the insurance end date (or date of the latest available record) and the event date was set to one day before the slack period. The variables were collected at the most recent time point of availability before the event date in both cohorts. This creates our desired imminent prediction paradigm, where a prediction of stroke reflects the imminent and overall likelihood for a future primary event. An illustration of this procedure is shown on Figure S1.

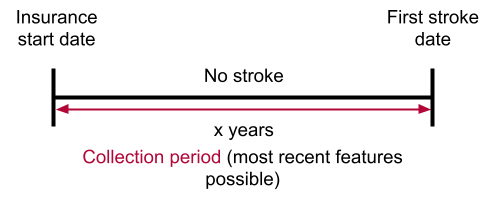

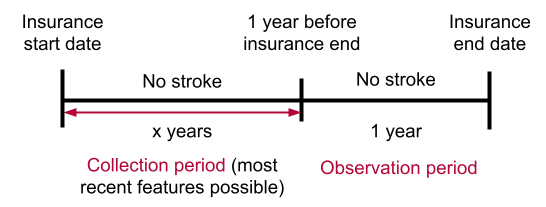

a)	b)

Figure S1: Illustration of event date identification strategy for stroke patients (a) and controls (b). In case of stroke patients, the event date was the date of the first registered stroke event. In case of controls, the event date was the date 1 year before the end date of their insurance. Variables were extracted from the most recent records of patients before the individual event dates.

Furthermore, the extracted, individual time periods and features were closely adjusted and matched between the cohorts. Firstly, to compensate for the 1 year shorter maximum insurance length of controls due to the slack period, the first insurance year of stroke patients was removed, in case they had the maximum insurance length. Secondly, the available insurance length was generally shorter for the stroke cohort, making matching by insurance length necessary to remove temporal bias. The insurance length of control patients was capped to resemble the same length in general as stroke patients to eliminate any temporal mismatch between the two cohorts. Lastly, an equal number of control patients were matched to the stroke cohort by age and insurance length. Consequently, mean age and insurance length were 76 and 3 respectively for both control and stroke cohorts.

#### Feature representation

We defined 4 different feature representation scenarios following 2 aspects: 1) Degree of ICD-10 granularity 2) Occurrence based (binary) or temporal representation of ICD-10 features.

The basis of the scenarios consists of the demographic features as well as care service levels. Care service features are given in days in any scenario as we have exact reports of when a service was taken. Scenario I includes ICD-10 disease codes as listed in Table 1. in a coarse fashion. In Scenario II, we exploited further subclasses of each ICD-10 class included in Scenario I. Subclasses are defined as the first 4 characters of the codes. Subclass variables with a lower prevalence than 5 percent were grouped or discarded by the same process outlined above. Next, to facilitate a distinction of high risk patients - i.e. high occurrence of many diseases - we infused temporal information about the duration of diseases. In Scenario III, we translated the original, binary ICD-10 classes by counting the days of the persistence of a variable since its first occurrence in the patient’s record. Similarly, in Scenario IV the same is done with the detailed subclasses. Table 2. Summarizes our defined feature scenarios.

|  |  | ICD-10 granularity | |
| --- | --- | --- | --- |
|  |  | Coarse | Detailed |
| Representation | Binary | Scenario I | Scenario II |
|  | Temporal | Scenario III | Scenario IV |

Table S2. Summary of feature representation scenarios

#### Model Development and Specification

For each defined feature set, traditional and machine learning models were trained and validated in the described nested cross-validation framework. The best model parameters were determined by grid-search and best validation performance across 4 folds. Fixed hyper parameters are shown in Table S3 and the defined parameter ranges for tuning in Table S4.

| **Model** | **Parameter** | **Range** |
| --- | --- | --- |
| **SVM** | Tolerance | 0.001 |
|  | Maximum iteration | 2500 |
| **NN** | Monitored measure | Validation accuracy |
|  | Mode | Max |
|  | Epochs | 50 |
|  | Minimum delta | 0.001 |
|  | Output activation | Sigmoid |
|  | Loss function | Binary cross entropy |
|  | Optimizer | Adam |
|  | Weight initialization | Uniform |
| Tree-boosting | OD Type | Iter |
|  | OD wait | 50 |

Table S3. Fixed hyper parameters for models, where not all parameters were tuned.

| **Model** | **Parameter** | **Range** |
| --- | --- | --- |
| **Lasso** | L1 regularisation | [0.01, 0.05, 0.1, 0.5, 1, 5, 10, 50, 100, 500] |
| **Elasticnet** | L1 regularisation | [0.0, 0.1, 0.25, 0.5, 0.75, 0.9] |
|  | L2 regularisation | [0.1, 0.25, 0.5, 0.75, 0.9] |
| **SVM** | C | [0.01, 0.1, 1, 3, 5] |
| **NN** | Batch size | [512, 2048] |
|  | Number of neurons | [[64, 128, 64], [128, 256, 128]] |
|  | Learning rate | [0.001, 0.0025] |
|  | Dropout rate | [0., 0.2] |
|  | Hidden activation | ['relu','elu'] |
| **Tree-boosting** | Depth | [4, 6, 8] |
|  | Leaf estimation iterations | [1, 4, 6] |
|  | Learning rate | [0.1, 0.2, 0.3] |

Table S3. Parameter ranges for each model, used for hyperparameter search implemented by grid-search.

#### Model performance

Results for all models with respect to all performance measures are shown in Table S4. Traditional models achieved the same AUROC in each scenario, while Lasso proved to be superior in terms of Sensitivity. The best penalty term was C=100.0 for split 1 and C=5.0 for split 2. Amongst ML models, Tree-boosting outperformed other models by far for all scenarios. The best parameters of the Tree-boosting method were depth = 6.0, leaf_estimation_iterations = 6.0 and learning_rate = 0.2 for both splits. Model calibration was assessed by the Brier score, Tree-boosting model seemed better calibrated with a Brier score of 0.12 in Scenario III.

| **Scenario** | **Performance measure** | **LR** | **Lasso** | **EN** | **SVM** | **NB** | **NN** | **TB** |
| --- | --- | --- | --- | --- | --- | --- | --- | --- |
| **Scenario I**  (Coarse ICD10) | **AUROC** | 0.61 | 0.61 | 0.61 | 0.57 | 0.59 | 0.64 | **0.75** |
|  | **SN** | 0.56 | 0.56 | 0.55 | 0.55 | 0.62 | 0.67 | **0.70** |
|  | **SP** | 0.59 | 0.59 | 0.60 | 0.60 | 0.50 | 0.52 | **0.64** |
|  | **BAcc** | 0.57 | 0.57 | 0.58 | 0.57 | 0.56 | 0.60 | **0.67** |
|  | **F1** | 0.57 | 0.57 | 0.56 | 0.56 | 0.59 | 0.62 | **0.67** |
|  | **Brier** | 0.24 | 0.24 | 0.24 | 0.43 | 0.31 | 0.23 | **0.20** |
| **Scenario II**  (Detailed ICD10) | **AUROC** | 0.62 | 0.62 | 0.62 | 0.58 | 0.58 | 0.64 | **0.75** |
|  | **SN** | 0.57 | 0.57 | 0.56 | 0.56 | 0.50 | 0.67 | **0.69** |
|  | **SP** | 0.59 | 0.59 | 0.60 | 0.60 | 0.62 | 0.51 | **0.64** |
|  | **BAcc** | 0.58 | 0.58 | 0.58 | 0.58 | 0.56 | 0.59 | **0.67** |
|  | **F1** | 0.58 | 0.58 | 0.57 | 0.57 | 0.53 | 0.62 | **0.68** |
|  | **Brier** | 0.24 | 0.24 | 0.24 | 0.42 | 0.37 | 0.23 | **0.20** |
| **Scenario III**  (Coarse ICD + Temporal information) | **AUROC** | 0.59 | 0.59 | 0.59 | 0.56 | 0.56 | 0.65 | **0.91** |
|  | **SN** | 0.59 | 0.58 | 0.54 | 0.58 | 0.36 | 0.70 | **0.84** |
|  | **SP** | 0.54 | 0.55 | 0.58 | 0.55 | 0.69 | 0.50 | **0.81** |
|  | **BAcc** | 0.56 | 0.56 | 0.56 | 0.56 | 0.52 | 0.60 | **0.83** |
|  | **F1** | 0.57 | 0.57 | 0.55 | 0.57 | 0.43 | 0.64 | **0.83** |
|  | **Brier** | 0.24 | 0.24 | 0.24 | 0.44 | 0.38 | 0.23 | **0.12** |
| **Scenario IV**  (Detailed ICD10 + Temporal information) | **AUROC** | 0.58 | 0.58 | 0.59 | 0.56 | 0.56 | 0.63 | **0.90** |
|  | **SN** | 0.59 | 0.59 | 0.57 | 0.58 | 0.32 | 0.69 | **0.83** |
|  | **SP** | 0.53 | 0.53 | 0.55 | 0.55 | 0.73 | 0.50 | **0.79** |
|  | **BAcc** | 0.56 | 0.56 | 0.56 | 0.56 | 0.53 | 0.59 | **0.81** |
|  | **F1** | 0.57 | 0.57 | 0.57 | 0.57 | 0.40 | 0.63 | **0.81** |
|  | **Brier** | 0.24 | 0.24 | 0.24 | 0.44 | 0.45 | 0.24 | **0.13** |

Table S4: Test performance of all models with coarse ICD-10 features (Scenario I), detailed ICD-10 features (Scenario II), temporal information about duration of coarse ICD-10 features (Scenario III) and of detailed ICD-10 features (Scenario IV). Values shown are calculated on test set. Bold depicts the best, blue the second best overall performance while red the best performance across traditional models per performance measure; AUROC - Area Under the ROC Curve, SN - Sensitivity, SP - Specificity, BAcc - Balanced Class Accuracy, F1 - F1 score, Brier - Brier score, LR - Logistic Regression, EN - ElasticNet, SVM - Support Vector Machine, NB - Näive Bayes, NN - Neural Network, TB - Tree-boosting.
